# Projecting national outpatient expenditure in South Korea: A scenario-based simulation of the impact of health literacy improvement

**DOI:** 10.1101/2025.11.19.25340606

**Authors:** Seokjun Moon, Junghun Yoo, Dunsol Go, Hocheol Lee

**Author notes:** Corresponding author: Hocheol Lee.

## Abstract

The South Korean government has set policy targets to increase the share of adults with sufficient health literacy to 70% by 2030. This study aimed to estimate changes in outpatient healthcare expenditure while maintaining the current health literacy distribution (status quo) and achieving the national health literacy target by 2030 (policy target). An autoregressive integrated moving average model was used to project outpatient expenditure from 2023 to 2030. The analysis incorporated the relative cost differences between sufficient and inadequate health literacy groups based on Korea Health Panel data from 2021. A calibration factor was applied to adjust for underestimation in the policy target scenario. The results revealed that by 2030, total outpatient expenditure is projected to reach KRW 105.6 trillion under the status quo and KRW 103.7 trillion under the policy target scenario, resulting in savings of approximately KRW 1.9 trillion (1.8%).

## Introduction

Health literacy (HL) refers to the ability to access, understand, appraise, and use health information to make informed decisions about healthcare, disease prevention, and health promotion, thereby supporting quality of life throughout the life cycle [1]. The global significance of HL has been increasingly emphasized. In the 2016 Shanghai Declaration, the World Health Organization (WHO) identified HL as one of three key health promotion strategies [2]. More recently, the revised Health System Performance Assessment Framework, released by the Organisation for Economic Co-operation and Development (OECD), highlighted the need for health systems to become more people-centered. In this context, the OECD underscored the importance of equipping populations with sufficient HL levels to support this shift [3].

Consistent with these global trends, South Korea has designated HL enhancement as a national policy objective. Health Plan 2030, announced in 2023, identifies HL improvement as a cores policy goal and sets the specific target of achieving a sufficient HL level among 70% of adults by 2030 [4].

Inadequate HL is significantly associated with increased mortality and morbidity rates, poor medication adherence, and low self-rated health status [5−7]. It is also associated with the increased use of unnecessary outpatient services, hospital admissions, and emergency department visits [1, 7–9]. Furthermore, it has been linked to higher expenditures related to outpatient and inpatient care, pharmaceutical use, and emergency services [9-13]. Inadequate HL not only leads to greater out-of-pocket spending at the individual level but also strains the healthcare system’s human and financial resources, thereby contributing to the national fiscal burden. Avoidable healthcare costs attributable to inadequate HL are estimated to account for approximately 3 to 8% of the total national health expenditure [10, 13–15].

South Korea reports the highest average number of outpatient visits among OECD countries, reaching three times the OCED average at 17.5 per capita in 2022 [16]. Their healthcare system is characterized by a privately dominated delivery system, no gatekeepers in primary care, and a fee-for-service payment model. These structural features collectively facilitate the frequent and often provider-driven use of services, thereby contributing to higher outpatient spending and partially explaining the exceptionally high levels of outpatient expenditure. Thus, improving HL may be particularly important because individuals with sufficient HL are more likely to make appropriate healthcare decisions and effectively navigate complex healthcare environments; this can reduce unnecessary outpatient usage and related expenditures [17–19].

However, empirical research on the association between HL and healthcare expenditure in Korea remains limited. Given the national policy goal of achieving a sufficient HL level among 70% of the population by 2030, investigating the relationship between HL and outpatient healthcare spending is timely and meaningful. Furthermore, projecting potential changes in outpatient expenditure under scenarios in which the HL policy target is achieved could provide valuable evidence for policy planning and resource allocation.

In this context, we aimed to project changes in outpatient healthcare expenditure in South Korea under a circumstance in which the national target of achieving sufficient HL among 70% of the population by 2030 is met.

## Materials and methods

### Study design

A scenario-based time series analysis was applied to estimate changes in outpatient healthcare expenditures resulting from improved HL among the South Korean population. Two scenarios were developed. First, the status quo scenario assumes that the population’s HL level will remain unchanged through 2030. Second, the policy target scenario assumes that 70% of the population will reach a sufficient HL level by 2030, aligning with national policy goals. The outpatient expenditure trends from 2023 to 2030 were projected for each scenario and then compared.

### Data source

#### National outpatient expenditure

Outpatient expenditure data from the Korea National Health Accounts, specifically item H.C.3 (outpatient expenditure), from 1970 to 2022 were used. This official statistical system was approved under Article 18 of the “Statistics Act of Korea” and is considered a timely, comprehensive source of information on national healthcare expenditure. The OECD, the WHO, and Eurostat jointly developed its framework, which has been widely adopted by numerous countries to produce standardized health expenditure accounts. The Korea National Health Accounts were formed based on the SHA 2011 guidelines and are intended to enhance users’ understanding of the national healthcare financing system [20]; they provide a comprehensive measure of total national healthcare expenditure, including public (e.g., insurer payments) and private spending (e.g., out-of-pocket expenses) as well as non-covered services.

#### Population

Population data were obtained from future population projections published by Statistics Korea from 1970 to 2030.

#### Parameters

This study utilized data from the 2021 Korea Health Panel (KHP), a nationally representative and government-approved statistical dataset targeting adults aged 19 years and older. The 2021 KHP includes a validated HL instrument, therefore, this study was limited to 2021 data, and a final sample of 8,630 individuals was used for the analysis.

Two key parameters were described. First, the relative difference in annual per capita outpatient healthcare expenditure by HL level was estimated. Outpatient costs were log-transformed and analyzed using an ordinary least squares regression model to address the right-skewed nature of the expenditure data and meet the regression assumptions. Furthermore, bootstrap analysis (100 iterations) with robust standard errors was used to enhance the precision of the coefficient estimates and confidence intervals. Based on this analysis, individuals with sufficient HL were assumed to spend 8.4% less on outpatient services than those with inadequate HL. The corresponding results are provided in Additional file 1. Second, the population distribution by HL level was calculated using descriptive statistics from the 2021 KHP. The analysis showed that 50.7% of the sample had inadequate HL and 49.3% had sufficient HL. Further details are presented in Additional file 2.

### Variables

#### Primary outcome

The primary outcome of this study was annual outpatient healthcare expenditure per capita (in KRW). Actual expenditure data from 1970 to 2022 were utilized to estimate future values from 2023 to 2030 using an autoregressive integrated moving average (ARIMA) model. Annual per capita expenditure was calculated by dividing the total outpatient expenditure by the projected population for each corresponding year.

#### Scenario modifier

HL was assessed using the Korean version of the HLS-EU-Q16, a 16-item instrument developed to capture individuals’ health-related competencies across three domains: healthcare (7 items), disease prevention (5 items), and health promotion (4 items). Each item was rated on a 5-point scale, ranging from “very difficult” to “very easy,” with an additional “don’t know” option. For scoring purposes, responses indicating difficulty (“very difficult” or “fairly difficult”) were coded as 0, and responses indicating ease (“fairly easy” or “very easy”) were coded as 1. This approach yielded a composite score ranging from 0 to 16.

It was categorized as inadequate (0–8), problematic (9–12), or sufficient (13–16).

However, in the context of multivariate analysis, several studies have employed binary classification, distinguishing HL levels as either inadequate (scores of 12 or below) or sufficient (scores above 12). This study adopted the latter approach, categorizing respondents into sufficient and inadequate HL groups based on this threshold [21−24].

#### Calibration factor

In the policy target scenario, outpatient healthcare expenditure for each HL group was estimated by applying a relative cost ratio to the national average per capita expenditure. This provided the HL-specific per capita costs, which we then multiplied by the projected population in each group to calculate total outpatient spending.

However, when these group-specific totals were added together, the sum was lower than the total projected expenditure in the baseline (status quo) scenario since individuals with sufficient HL are expected to use less care, thereby lowering the overall estimate. A calibration factor was applied to correct this gap. This factor was calculated as the ratio of the totals in the status quo and policy target scenarios. The HL-specific per capita costs were then multiplied by this factor to adjust them upward, ensuring that the total matched the baseline. The adjusted per capita costs were multiplied by the population again to produce the final expenditure estimates, as follows:

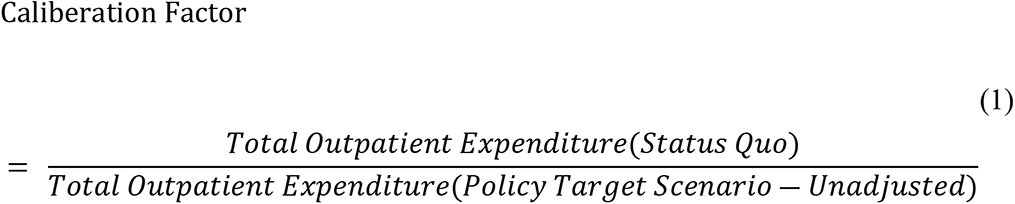

### Statistical modeling and scenario simulation

#### Status quo scenario

Outpatient healthcare expenditures from 2023 to 2030 were projected using historical data from the Korea National Health Accounts from 1970 to 2022. An ARIMA (p, d, q) model was used to incorporate three components: autoregression (AR), differencing (I), and moving average (MA). To avoid overfitting or underfitting, appropriate values must be determined for each parameter. First, the differencing order (d) was identified by testing for stationarity using a unit root test. Then, the autoregressive order (p) and moving average order (q) were selected based on the analysis of the autocorrelation function and partial autocorrelation function plots. The model was validated and selected based on the lowest Akaike Information Criterion (AIC) value, which indicates the best fit among competing specifications.

#### Policy target scenario

This analysis estimated changes in outpatient healthcare expenditure under the policy target scenario in which 70% of the population achieves sufficient HL by 2030, following the methodological approaches used in previous studies [25]. First, the annual per capita outpatient healthcare expenditure from 2023 to 2030 was estimated. The projected total outpatient expenditure for each year was divided by the projected population to obtain per capita values.

Second, outpatient expenditure per capita by HL level was calculated. The relative difference in outpatient spending between HL groups was derived from a sub-analysis of the 2021 KHP data. Based on prior findings indicating that individuals with sufficient HL spend 8.4% less on outpatient care annually than those with inadequate HL, this relative difference was applied to the national average per capita expenditure. For example, the per capita expenditure for the sufficient HL group was estimated to be 91.6% of the national average, whereas that for the inadequate HL group was assumed to be at the baseline level.

Third, the population distribution by HL level was projected for each year from 2023 to 2030. The baseline distribution in 2021 was estimated to be 49.3% and 50.7% for sufficient and inadequate HL, respectively. The proportion of individuals with sufficient HL was assumed to increase linearly to 70% by 2030, which is in line with the national target.

Fourth, total outpatient expenditure under the policy target scenario was computed by multiplying HL-specific per capita expenditure by the corresponding HL-specific population for each year. However, the direct application of the relative spending difference may result in a discrepancy between the total policy target scenario estimate and baseline projection. Therefore, a calibration factor was introduced to address this issue. This factor was calculated as the ratio of total outpatient expenditure under the status quo scenario to that under the unadjusted policy target scenario. The HL-specific per capita expenditure was then adjusted using this factor and the total expenditure was recalculated accordingly. Finally, the projected outcomes under the status quo and policy target scenarios were compared to assess the impact of improved population HL.

#### Ethics Statement

This study used fully anonymized secondary data from the Korea Health Panel Survey (KHPS). The KHPS data collection was approved by the Institutional Review Board of the Korea Institute for Health and Social Affairs (KIHASA) (IRB No. 2022-017), and all personal identifiers were removed before the dataset was released for research use. The analyses in this manuscript were conducted as part of my doctoral dissertation and received an exemption from further review from the Yonsei University Mirae Institutional Review Board (IRB No. 1041849-202409-SB-202-01). Because the dataset was fully anonymized and publicly accessible, the requirement for informed consent was waived. No identifiable information was available to the authors at any stage of the research.

## Results

A unit root test was conducted to assess the stationarity of the time series data and determine the ARIMA (p, d, q) model. This test confirmed the absence of a unit root, and first-order differencing (d = 1) was applied. Among the candidate models with d = 1, the ARIMA (1, 1, 3) model with the lowest AIC value was selected and used to project outpatient healthcare expenditure from 2023 to 2030.

### Status quo scenario

Based on data from the Korea National Health Accounts, actual outpatient healthcare expenditures amounted to KRW 60.3 trillion in 2022 and were projected to increase to KRW 63.5 trillion in 2023 and KRW 105.6 trillion by 2030. Per capita outpatient expenditure was KRW 1,166,359 in 2022 and projected to reach KRW 1,228,380 in 2023 and KRW 2,059,014 by 2030, corresponding to an average annual growth rate of 7.4% (Table 1, Fig 1).

**Fig 1.**
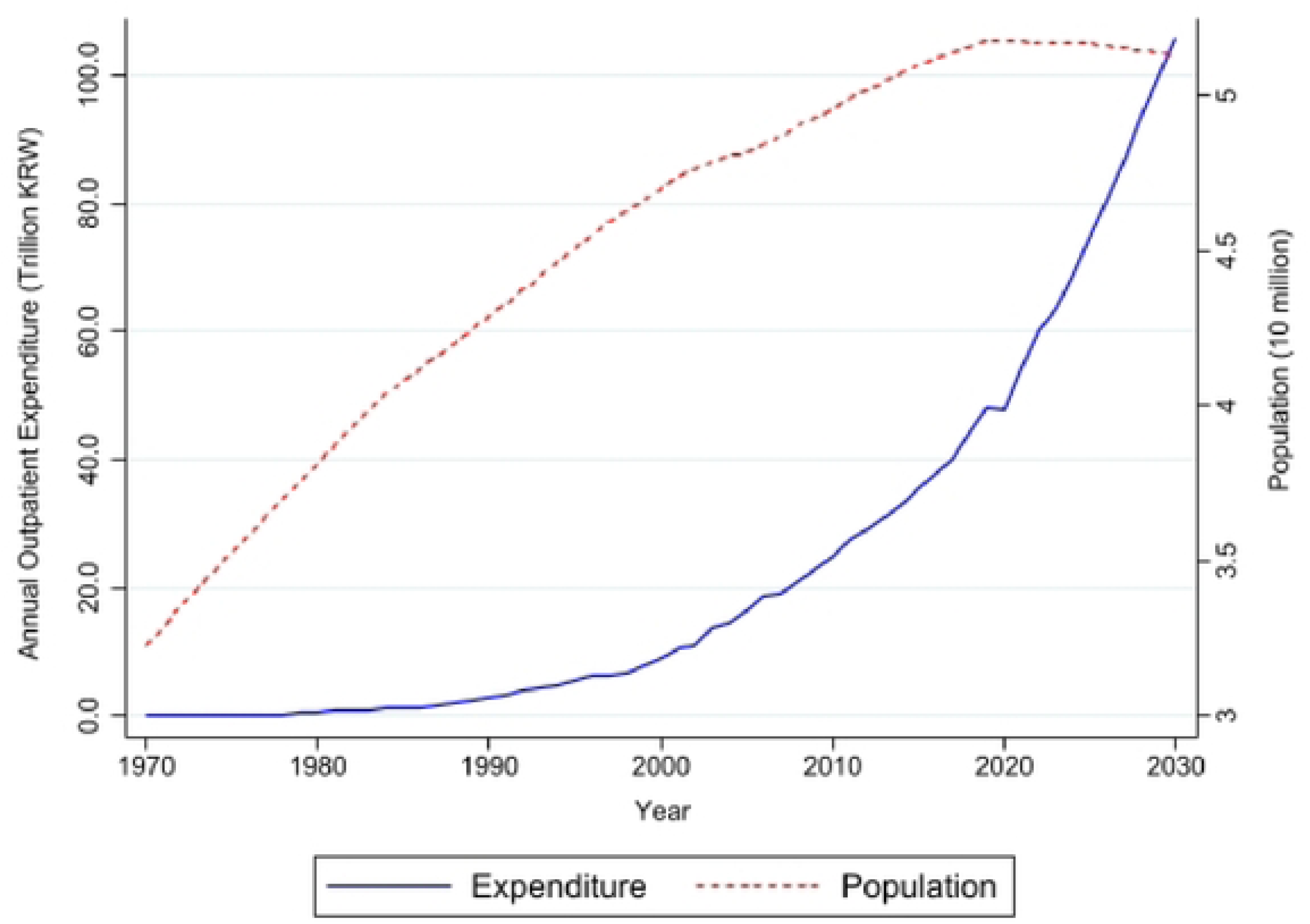
Annual outpatient spending and population projection (1970–2030)

**Table 1.**
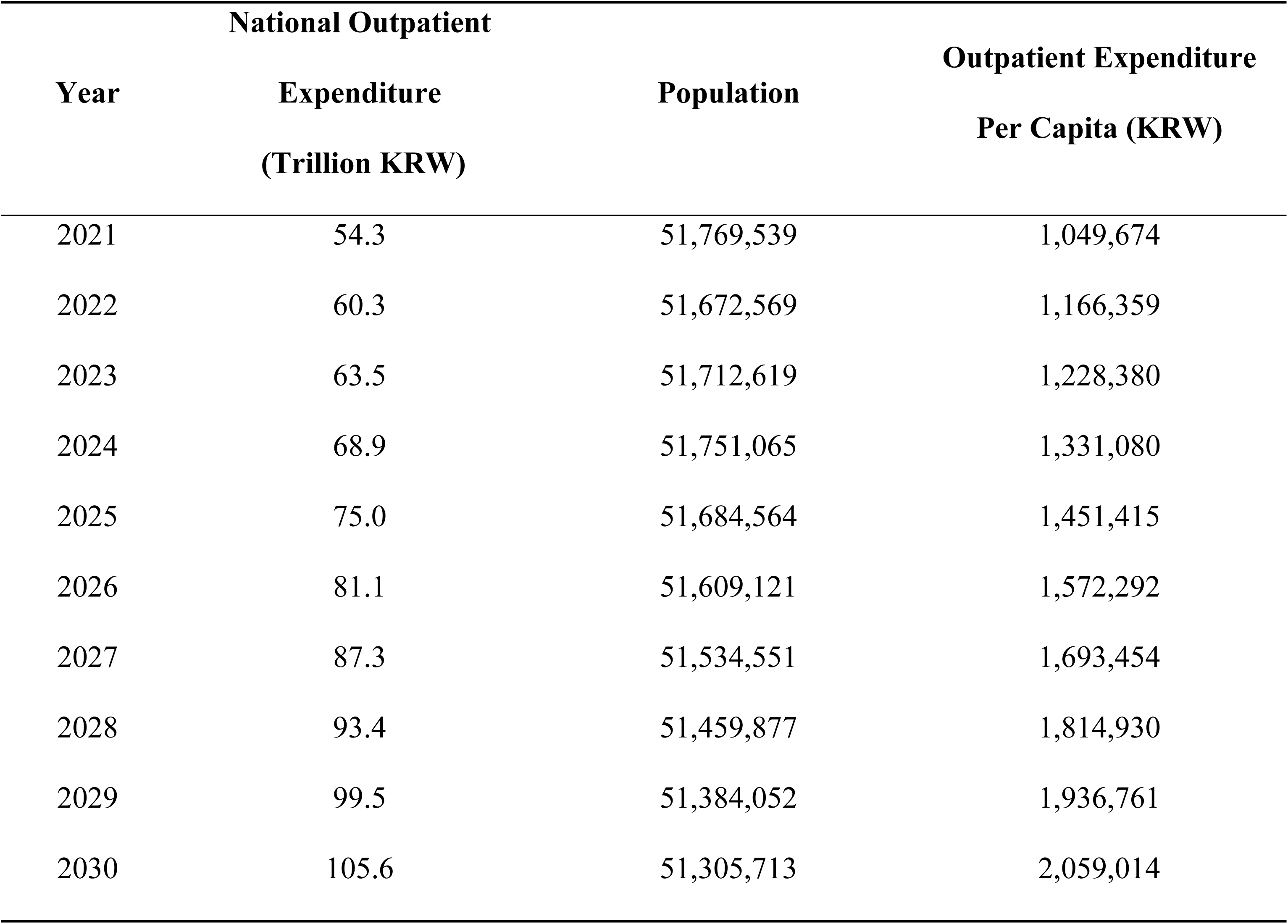
Status Quo Scenario: Outpatient Expenditure (1970–2030)

### Policy target scenario

The estimated per capita outpatient healthcare expenditure by HL level in 2022 was KRW 1,068,385 for individuals with sufficient HL and KRW 1,166,350 for those with inadequate HL. The projections indicated that figures would increase to KRW 1,125,196 and 1,228,380, respectively, by 2023, and to KRW 1,886,056 and 2,059,014, respectively, by 2030.

When total outpatient expenditure was calculated by multiplying the per capita expenditure by the corresponding population for each HL level by year, the result was underestimated compared with the original projection. To correct for this discrepancy, a calibration factor of 1.043 was derived, representing the ratio between the original estimate and the underestimated total.

After applying this calibration factor, the adjusted per capita outpatient expenditure for 2023 was estimated to be KRW 1,173,806 and 1,281,447 for the sufficient and inadequate HL groups, respectively. These values were projected to rise to KRW 1,967,536 and 2,147,965, respectively, by 2030 (Table 2).

**Table 2.**
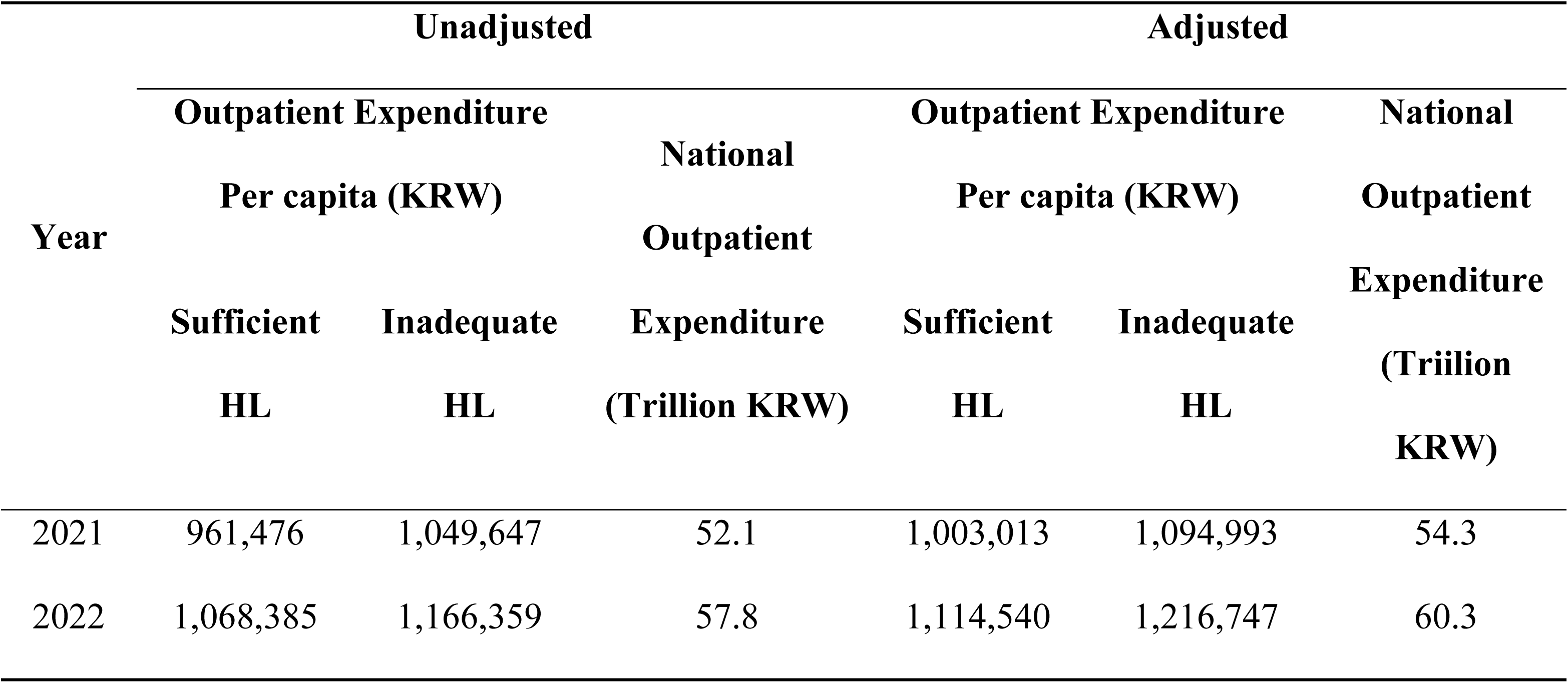
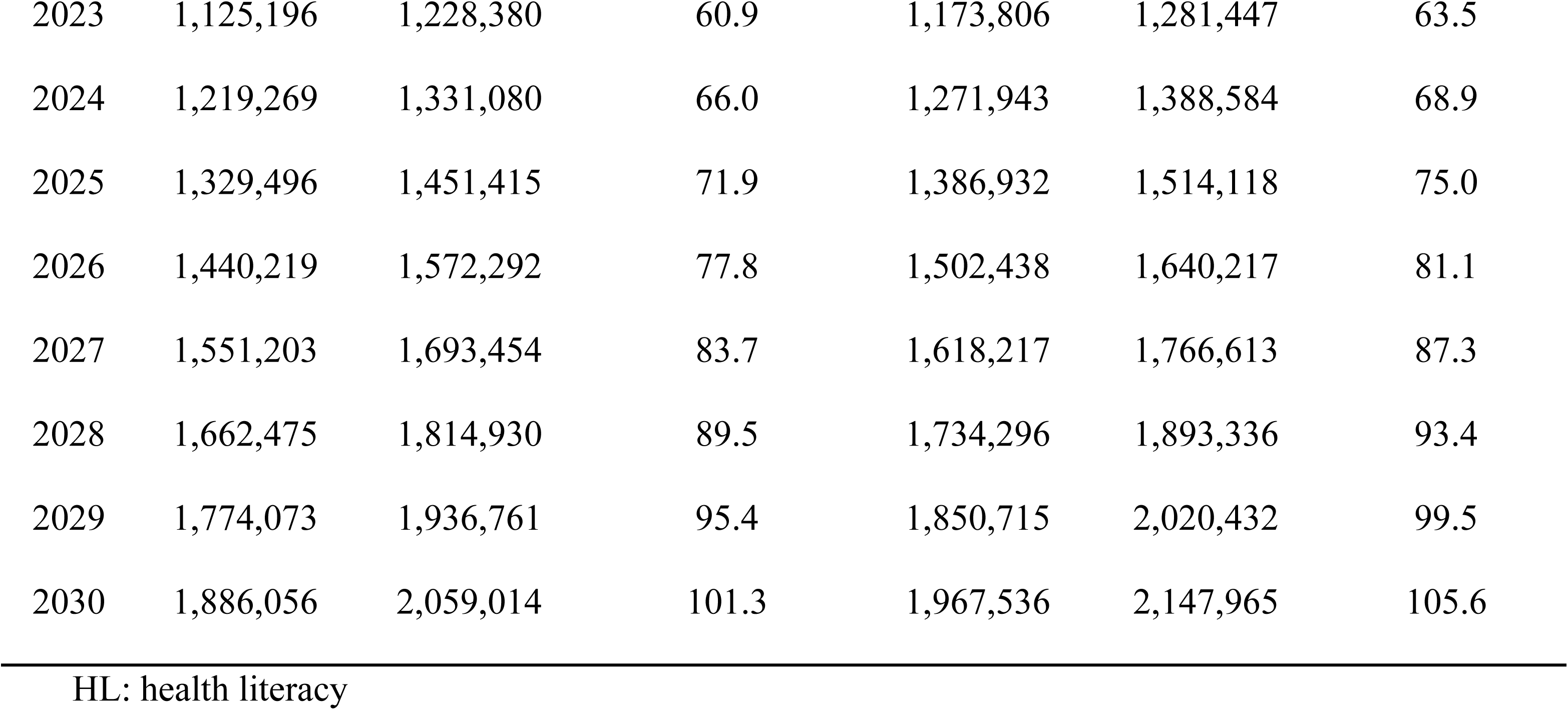
Per Capita Outpatient Expenditure by Health Literacy Level After Applying the Calibration Factor.

| Year | Unadjusted |  |  | Adjusted |  |  |
| --- | --- | --- | --- | --- | --- | --- |
|  | Outpatient Expenditure |  | National<br>Outpatient<br>Expenditure<br>(Trillion KRW) | Outpatient Expenditure |  | National<br>Outpatient<br>Expenditure<br>(Trillion KRW) |
|  | Per capita (KRW) |  |  | Per capita (KRW) |  |  |
|  | Sufficient | Inadequate |  | Sufficient | Inadequate |  |
|  | HL | HL |  | HL | HL |  |
| 2021 | 961,476 | 1,049,647 | 52.1 | 1,003,013 | 1,094,993 | 54.3 |
| 2022 | 1,068,385 | 1,166,359 | 57.8 | 1,114,540 | 1,216,747 | 60.3 |
| 2023 | 1,125,196 | 1,228,380 | 60.9 | 1,173,806 | 1,281,447 | 63.5 |
| 2024 | 1,219,269 | 1,331,080 | 66.0 | 1,271,943 | 1,388,584 | 68.9 |
| 2025 | 1,329,496 | 1,451,415 | 71.9 | 1,386,932 | 1,514,118 | 75.0 |
| 2026 | 1,440,219 | 1,572,292 | 77.8 | 1,502,438 | 1,640,217 | 81.1 |
| 2027 | 1,551,203 | 1,693,454 | 83.7 | 1,618,217 | 1,766,613 | 87.3 |
| 2028 | 1,662,475 | 1,814,930 | 89.5 | 1,734,296 | 1,893,336 | 93.4 |
| 2029 | 1,774,073 | 1,936,761 | 95.4 | 1,850,715 | 2,020,432 | 99.5 |
| 2030 | 1,886,056 | 2,059,014 | 101.3 | 1,967,536 | 2,147,965 | 105.6 |

HL: health literacy

Based on 2023 estimates, multiplying the number of individuals with sufficient HL (27,873,102 people, 53.9%) by their per capita outpatient expenditure (KRW 1,125,196), and adding this to the product of the number of individuals with inadequate HL (23,839,517 people, 46.1%) and their corresponding per capita expenditure (KRW 1,281,447), the total annual outpatient healthcare expenditure was approximately KRW 63.5 trillion.

Assuming that the proportion of individuals with sufficient HL will increase to 70.0% by 2030, the total annual outpatient expenditure was projected by multiplying 35,913,999 individuals (70.0%) by KRW 1,967,536 and 15,391,714 individuals (30.0%) by KRW 2,147,965. The resulting combined expenditure was approximately KRW 103.7 trillion (Table 3).

**Table 3.** Policy Target Scenario: Outpatient Expenditure (2023–2030)

| Year | Proportion of<br>Sufficient HL<br>(%) | Population |  | Outpatient Expenditure<br>Per capita (KRW) |  | National<br>Outpatient<br>Expenditure<br>(Trillion KRW)<br>(A*C) + (B*D) |
| --- | --- | --- | --- | --- | --- | --- |
|  |  | Sufficient | Inadequate | Sufficient | Inadequate |  |
|  |  | HL | HL | HL | HL |  |
|  |  | A | B | C | D |  |
| 2021 | 49.3 | 25,522,383 | 26,247,156 | 1,003,013 | 1,094,993 | 54.3 |
| 2022 | 51.6 | 26,663,046 | 25,009,523 | 1,114,540 | 1,216,747 | 60.1 |
| 2023 | 53.9 | 27,873,102 | 23,839,517 | 1,173,806 | 1,281,447 | 63.3 |
| 2024 | 56.2 | 29,084,099 | 22,666,966 | 1,271,943 | 1,388,584 | 68.5 |
| 2025 | 58.5 | 30,235,470 | 21,449,094 | 1,386,932 | 1,514,118 | 74.4 |
| 2026 | 60.8 | 31,378,346 | 20,230,775 | 1,502,438 | 1,640,217 | 80.3 |
| 2027 | 63.1 | 32,518,302 | 19,016,249 | 1,618,217 | 1,766,613 | 86.2 |
| 2028 | 65.4 | 33,654,760 | 17,805,117 | 1,734,296 | 1,893,336 | 92.1 |
| 2029 | 67.7 | 34,787,003 | 16,597,049 | 1,850,715 | 2,020,432 | 97.9 |
| 2030 | 70.0 | 35,913,999 | 15,391,714 | 1,967,536 | 2,147,965 | 103.7 |

HL: health literacy

The projected 2030 outpatient healthcare expenditure under the policy target scenario was estimated at KRW 103.7 trillion, compared to KRW 105.6 trillion under the status quo scenario. The resulting difference of KRW 1.9 trillion represents the estimated impact of achieving the national HL goal, corresponding to approximately 1.8% of the total expenditure under the status quo scenario (Table 4, Fig 2).

**Fig 2.**
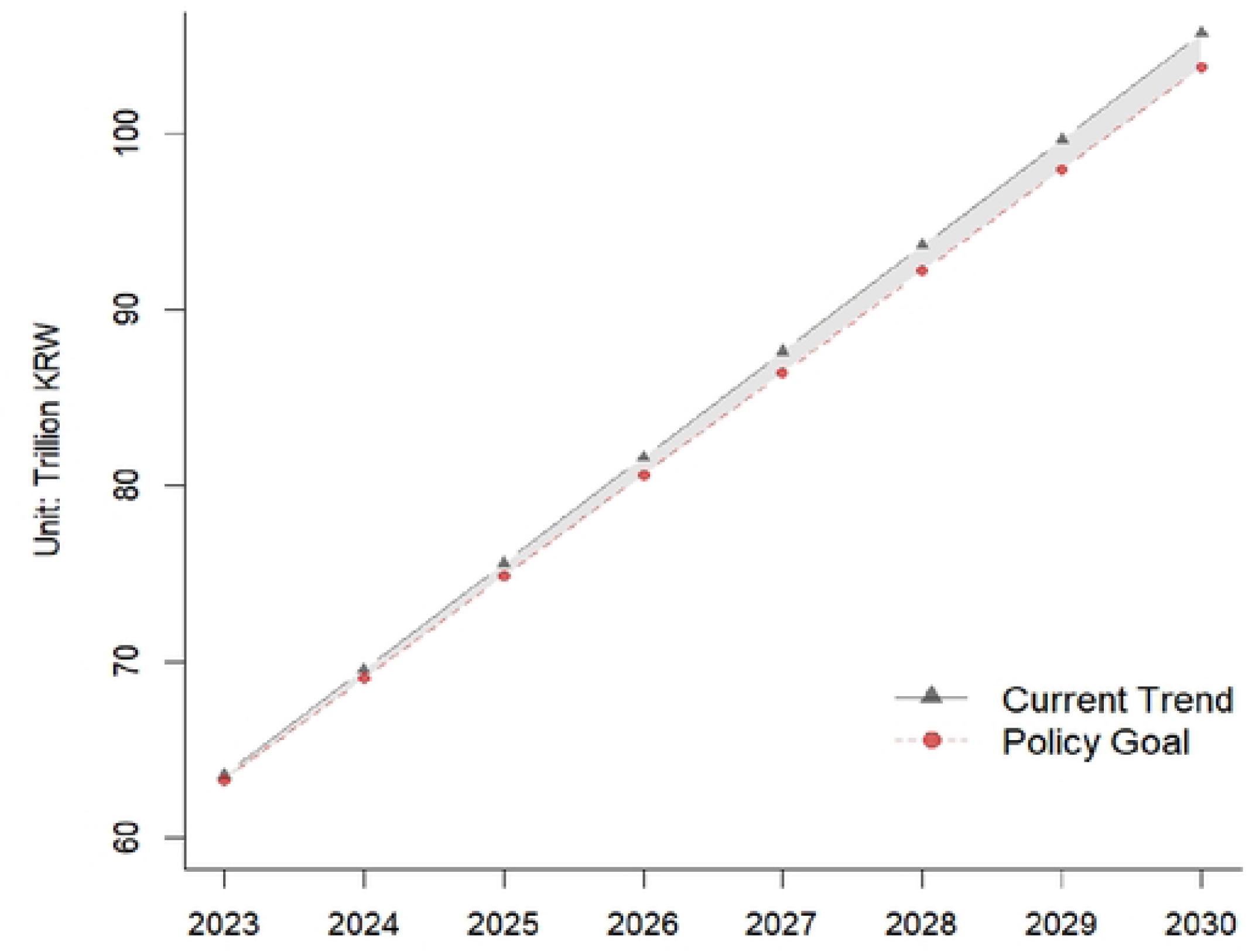
Comparing the status quo and policy target scenarios.

**Table 4.** Comparing Scenarios.

| Year | National Outpatient Expenditure |  | Difference (A-B) |
| --- | --- | --- | --- |
|  | (Trillion KRW) |  |  |
|  | Status Quo (A) | Policy Target (B) |  |
| 2023 | 63.5 | 63.3 | 0.2 |
| 2024 | 68.9 | 68.5 | 0.4 |
| 2025 | 75.0 | 74.4 | 0.6 |
| 2026 | 81.1 | 80.3 | 0.8 |
| 2027 | 87.3 | 86.2 | 1.1 |
| 2028 | 93.4 | 92.1 | 1.3 |
| 2029 | 99.5 | 97.9 | 1.6 |
| 2030 | 105.6 | 103.7 | 1.9 |

## Discussion

This study examined changes in outpatient healthcare expenditures according to the level of HL in the Korean population. An ARIMA (p, d, q) model was used for the time series analysis. After confirming stationarity through first-order differencing and comparing AIC values across models, ARIMA (1,1,3) was selected as the optimal model. Based on this model, outpatient expenditure under the status quo scenario was projected to increase to KRW 105.6 trillion by 2030, with per capita spending reaching KRW 2,059,014. When estimating expenditures by HL level, individuals with sufficient HL spent KRW 961,476 per capita in 2021, whereas those with inadequate HL spent KRW 1,049,647. These figures were projected to increase to KRW 1,967,536 and 2,147,965, respectively, by 2030. In the policy target scenario, the proportion of the population with sufficient HL was assumed to increase from 49.3% by 2021 to 70% by 2030. Under this assumption, the total outpatient expenditure in 2030 was estimated to be KRW 103.7 trillion—1.9 trillion (or 1.8%) lower than the estimate in the status quo scenario.

These findings suggest that achieving the national HL target could yield economic benefits of approximately KRW 2 trillion annually for outpatient care alone. Prior international studies have estimated that 3–5% of total healthcare spending can be attributed to inefficient use resulting from inadequate HL. In the Korean context, even a 1–2% annual reduction in outpatient spending would mitigate rising healthcare costs and enhance the financial sustainability of the healthcare system [13, 14].

Moreover, improved HL may not only reduce direct medical costs (e.g., treatment and medication expenses) but also lower indirect costs (e.g., productivity loss and caregiver burden) [26]. These effects may be even more pronounced among older populations, further supporting the need to invest in HL improvement from a long-term healthcare efficiency perspective.

Although Korea’s current national health promotion plan aims to increase the share of adults with sufficient HL to 70%, policy discussions have largely focused on HL as a tool for health promotion rather than a lever for healthcare system efficiency. By contrast, several countries have adopted more strategic approaches, framing HL improvement as a cost-effective health system reform measure [13, 26–28]. For example, the United States has recognized inadequate HL as a key driver of inefficiency and responded by introducing a national action plan to improve HL as part of broader efforts to reduce healthcare costs and improve outcomes [29].

Korea should adopt similar steps to integrate the HL policy more fully into its broader health system and fiscal strategy [26, 30, 31]. When HL is addressed using a comprehensive, system-level approach, the country stands to gain not only improved health outcomes but also greater equity and more efficient use of limited healthcare resources.

This study had several limitations. First, the ARIMA model does not consider demographic changes or external shocks. Given Korea’s rapidly aging population and potential policy reforms, projections based solely on historical trends may underestimate future expenditure and overlook systemic shifts. Second, we assumed a linear increase in sufficient HL from 2021 to 2030. However, no national data currently track HL trends over time, and this assumption is based on a single cross-sectional dataset. The trajectory of HL improvement remains uncertain without longitudinal monitoring. Third, sufficient HL was defined using HLS-EU-Q16 scores from the 2021 KHP. Although Korea has not yet implemented a national HL indicator, a monitoring tool is being developed based on the same instrument, considering sensitivity, specificity, and threshold-based criteria [32]. Finally, the 8.4% cost differential between HL groups was drawn from preliminary dissertation findings. Therefore, this estimate should be validated by using more representative longitudinal data.

## Conclusions

This study used a scenario-based ARIMA time series model to project outpatient healthcare expenditure in South Korea under two scenarios: maintaining the status quo and achieving the national HL policy target. The findings suggest that increasing the proportion of the population with sufficient HL to 70% by 2030 could reduce national outpatient expenditure by approximately KRW 1.9 trillion annually, or 1.8% of the projected spending under the status quo. These results underscore the potential of HL not only as a health promotion tool but also as a strategic lever for improving healthcare system efficiency and financial sustainability. Given Korea’s rapidly aging population and structurally high outpatient utilization rate, investing in HL improvement could yield both individual and systemic benefits. Policymakers should view HL as a core element of health system reform and incorporate it into broader strategies for demand management, health equity, and long-term fiscal sustainability.

## Data Availability

All relevant data are within the manuscript and its Supporting Information files.

## Acknowledgments

No acknowledgments to disclose.

## Availability of data and materials

- National Health Accounts (outpatient healthcare expenditure): Available from Statistics Korea at https://kosis.kr/index/index.do.
- Population projections: Available from Statistics Korea at https://kosis.kr/index/index.do.
- Korea Health Panel (KHP): Available from the Korea Health Panel Study (https://www.khp.re.kr/) upon application and approval.

## Supporting Information

**Additional file 1. Multivariate Analysis: Impact of Health Literacy on Outpatient Expenditure**

**Additional file 2. Descriptive Analysis on Korea Health Panel 2021**

- Description of data

These data are derived from the author’s doctoral dissertation and are included in a related manuscript currently under review. They are presented here as background information only. Citation details will be updated upon publication, if applicable.

